# The Impact of Electronic Order Communications on Laboratory Turnaround Times in Acute Hospital Care

**DOI:** 10.1101/2024.01.06.24300924

**Authors:** S. Gallier, X. Zou, F. Evison, J. Hodson, J. Atia, C Webster, M. Garrick, J. Coleman, T. Pankhurst, S. Ball, K. Nirantharakumar, E. Sapey

## Abstract

**Objective:** To examine the impact of computerised physician order entry (CPOE) systems upon laboratory turnaround times (LTAT), namely the time from recording the collection of a blood sample to the results being reported, within a large acute hospital.

**Materials and methods:** 1,810,311 blood samples taken between 1^st^ January 2014 and 31^st^ December 2019 were included. Changes in LTAT over the 24 months pre- and 18 months post-CPOE implementation were analysed using a segmented regression approach. The primary analysis assessed the median LTAT across the whole hospital, with secondary analyses assessing subgroups defined by clinical settings.

**Results:** CPOE implementation was associated with a step-change reduction in the median LTAT of 31.7 minutes (95% CI: 25.5-37.9, p<0.001). This was sustained over eighteen months post-implementation of CPOE despite the number of samples increasing by an average of 31% in this post-implementation period. Analysis by broad clinical specialty found all subgroups of wards considered to have a significant reduction in LTAT post-CPOE, either in the form of a step-change reduction, or an increasing rate of change.

**Discussion and Conclusion:** The implementation of CPOE within an acute hospital improves the average LTAT over a prolonged period, despite an increase in the number of samples collected. This could improve care efficiencies. Understanding the likely reduction in LTAT also provides information to support an economic evaluation of the implementation of such a system into a new setting.

## BACKGROUND

With global health systems under significant pressure, there is considerable interest in increasing the productivity and efficiency of healthcare services. Clinical laboratory services (such as Biochemistry, Haematology, and Immunology) are a vital component of healthcare provision, with up to 70-80% of all healthcare decisions affecting diagnosis or treatment involving a laboratory investigation.^(1)^ Laboratory test results can facilitate decisions to discharge, treat and admit, including escalation of care decisions. Consequently, the laboratory turnaround time (LTAT) is an important marker of a laboratory service, and is often used as a key performance indicator in healthcare settings.^(2-4)^ LTAT is usually defined as the time from receiving a specific test request to reporting the result.^(5)^

Digital healthcare technologies have been promoted as a means to improve the efficiency of care, but with recognition that robust clinical validation is necessary prior to widespread adoption.^(6)^ Computerised Provider Order Entry (CPOE) systems are computer-assisted systems that replace a hospital’s paper-based ordering system by automating medication, test, or sample ordering and reporting processes.^(7)^ Studies assessing the impact of CPOE on medication prescriptions have consistently shown improvements in error rates and adverse drug interactions.^(8-13)^ The impact of CPOE on operational processes has been assessed in a limited number of studies, with a focus on medicine dispensing.^(14, 15)^ However, the impact of CPOE on LTAT across a whole hospital system over time and the potential operational efficiency gains have not yet been assessed.

University Hospitals Birmingham NHS Foundation Trust (UHB) is one of the largest NHS Trusts in the United Kingdom, providing direct acute services and specialist care across four hospital sites including the Queen Elizabeth Hospital Birmingham (QEHB). QEHB uses an Electronic Health Record (EHR) called Prescribing Information and Communications System (PICS), a rules-based prescription-support system that includes clinical documentation; all physiological and laboratory measurements; provides real-time drug prescribing checks and recommendations; as well as supporting institutional and individual audit of healthcare processes.

## OBJECTIVE

This study aimed to assess the impact of the CPOE system on LTAT for processing blood tests within QEHB. Trends were assessed over the 24 months pre- and 18 months post-CPOE implementation for each ward. The primary analysis was performed using data pooled across all wards, with subgroup analyses performed within different clinical specialties. As a sensitivity analysis, all analyses were repeated for the period of LTAT within the laboratory section of the pathway.

## METHODS

The study was supported by PIONEER, a Health Data Research Hub in Acute Care. Ethical approvals for the study were provided by the East Midlands – Derby REC (reference: 20/EM/0158).

### Laboratory processing and details of CPOE

The laboratory reporting cycle at QEHB prior to and after the implementation of CPOE is summarised in **Figure 1**, which describes time stamped points included in the current analysis and how the sensitivity analysis was conducted with **Figure 1a** representing processes pre-CPOE and **Figure 1b** post-CPOE implementation.

**Figure 1.**
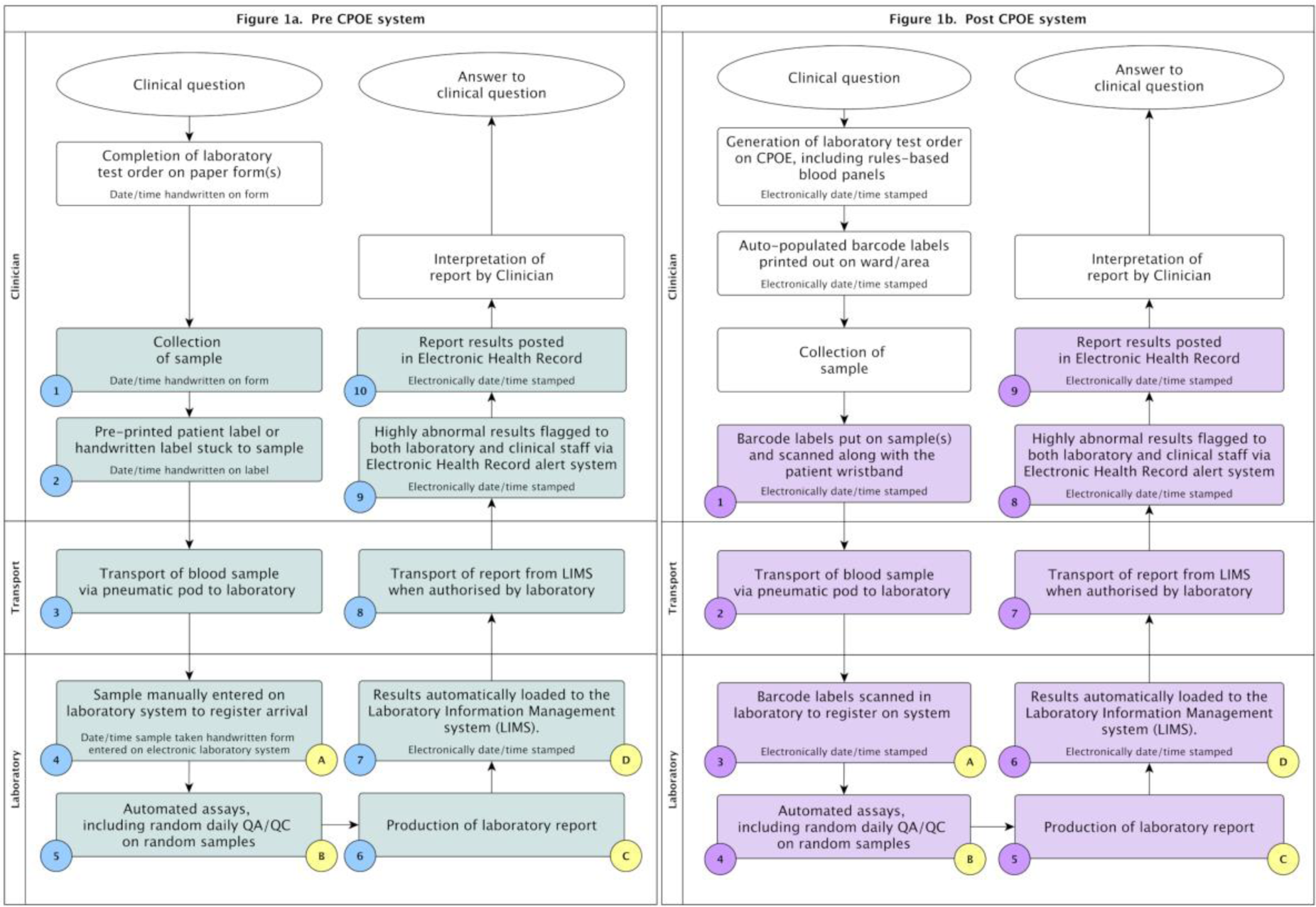
Request report cycle showing the process flows for blood tests ordered at QEHB pre- and post-CPOE. **Legend**. The figure illustrates all steps in the process flow of blood test ordering, including the forming of a clinical question; sample collection and transport; laboratory analysis; and the reporting, interpretation and decision making from the results. The steps comprising the period of LTAT are highlighted in blue for the pre-CPOE process (Figure A) and in purple for the post-CPOE process (Figure B). In each case, the steps included in the sensitivity analysis of Lab-LTAT are indicated with yellow circles. CPOE – Computerised Physician Order Entry; LIMS – Laboratory Information Management System; LTAT – Laboratory Turnaround Time; QA/QC: Quality Assurance/Quality Control.

CPOE was implemented across the hospital in a stepwise fashion from 21^st^ January 2016 to 5^th^ March 2019. CPOE introduced a range of changes to the process of recording the flow of samples through the hospital, as described in **Figure 1b**. Specifically, the introduction of wristbands with barcodes, electronic blood order forms, rules-based blood panels and printed labels. Other than the implementation of CPOE there were no other major changes in laboratory processing (e.g., measuring assays, reporting, quality assurance or quality control protocols) during the study period.

### Study population

All in-patients attending QEHB between 1^st^ January 2014 and 31^st^ December 2019 who had blood samples taken on the premises were initially considered for inclusion in the study population. All blood samples collected were initially considered for inclusion in the study, with no limit applied to the number of samples per admission, leading to a total of 4,422,971 samples.

Patients were then excluded if: i) they were admitted for dialysis or ii) the bloods were sent to external laboratories for analysis. After applying these exclusions, initial assessment of the distribution of LTAT for the remaining samples identified some extreme outliers. The majority were tests ordered without sample collection or complex tests requiring extended analysis. As neither of these situations represented a typical testing process, outliers with LTAT of more than seven days were excluded from the analysis. **See Supplementary** Figure 1.

In order to maintain a consistent cohort of wards, only the samples collected in the period from the 24 months pre-CPOE and 18 months post-CPOE for each ward were considered in the analysis of LTAT. Wards where CPOE was implemented less than 18 months from the date of data collection were excluded. After exclusions, a total of 1,810,311 blood samples contributed to the analysis of LTAT.

### Data capture

PICS has a comprehensive, time stamped audit database of all actions taken within the system. Data on each laboratory sample request during admissions, including request generation and report dates and times, ward, and requesting specialty, were extracted to calculate the LTAT. Prior to the implementation of CPOE, LTAT was defined as the time from the sample being collected on the handwritten label to the results being reported on the EHR (see ***Figure 1a***). After CPOE implementation, LTAT was defined as the time from the barcodes on the sample label being scanned on the ward to the results being reported on the EHR (see ***Figure 1b***). Where the same blood sample was used for multiple tests, LTAT was defined as first result to be reported (i.e. the shortest LTAT for each sample). In addition to the overall LTAT, the laboratory-specific LTAT (Lab-LTAT) was also assessed as a sensitivity analysis, which was defined as the time from a sample being registered on the system after arrival at the laboratory to the results being reported on the EHR (see yellow steps A-D in ***Figure 1a/b***).

For the primary analysis, the median LTAT was calculated across all samples collected within monthly intervals. Since the date of CPOE implementation varied by ward, the month numbers were standardised based on the date that CPOE was introduced on the ward. For example, CPOE was introduced to the first ward on 21^st^ January 2016; hence, “Month 0” for this ward would include samples collected between 21^st^ January 2016 and 20^th^ February 2016, with previous months assigned negative values, and subsequent months assigned positive values. For subgroup analyses, medians were calculated similarly for individual wards, or groups of wards within a specialty.

### Analysis

Trends over time in the monthly median LTAT, and the impact of CPOE were analysed using a segmented linear regression of interrupted time series (ITS) approach.^(16)^ A linear regression model was used, with the median monthly LTAT as the dependent variable and three covariates, which quantified the pre-CPOE gradient, the step-change occurring immediately post-CPOE, and the change in the gradient between the pre- and post-CPOE periods, respectively. A similar approach was also used to assess the changes in the monthly numbers of completed samples over the study period.

All analyses were performed using IBM SPSS 22 (IBM Corp. Armonk, NY) or R (R Core team, 2020), with p<0.05 deemed to be indicative of statistical significance throughout.

## RESULTS

### Numbers of samples

CPOE was introduced to the first two wards on 21^st^ January 2016, with five wards having CPOE by the end of 2016, 15 by the end of 2017, 37 by the end of 2018, and all 39 wards being live by March 2019. The numbers of blood order samples per month was stable over the two years prior to CPOE being introduced (p=0.160), with approximately 53,000 samples being processed per month (see **Figure 2A**). However, after CPOE roll out, the number of samples per month began to increase progressively (p<0.001). Due to the staggered implementation of CPOE across the wards, it was difficult to identify the direct impact on CPOE on the numbers of bloods samples requested. A subsequent analysis considered the N=27 wards with at least 18 months of follow-up post-CPOE implementation, with the calendar months being centred on the date of CPOE implementation at a ward. Here, there was a step-change increase in the number of samples processed immediately after CPOE implementation, with an average increase of 12,541 samples per month (95% CI: 11,481 – 13,602, p<0.001), representing a 31% increase (***Figure 2B***). This higher number of samples was then sustained over the 18 months post-CPOE implementation.

**Figure 2.**
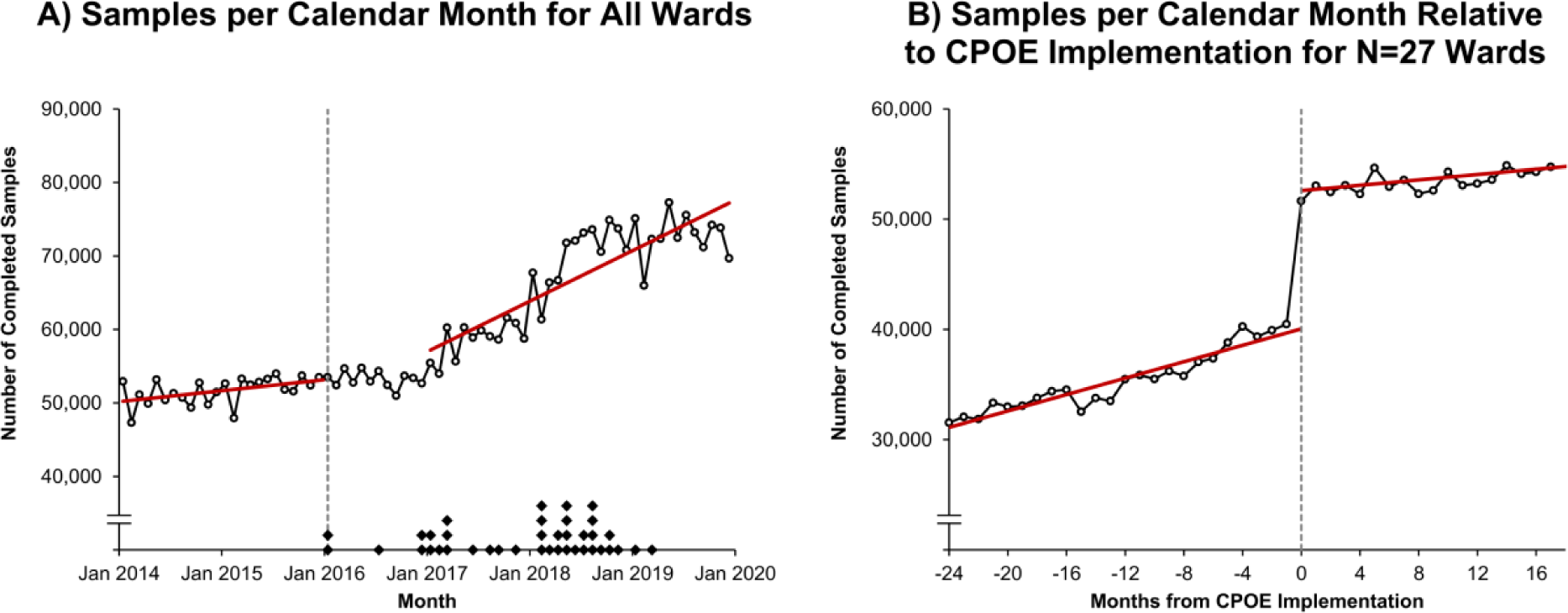
Monthly numbers of completed blood sample orders for inpatients at QEHB. **Legend.** In Figure A, points represent the total numbers of completed blood samples within each calendar month and are plotted at the mid-point of the month. Trend lines are from a segmented regression model; this excluded data for the year after CPOE implementation commenced (i.e., 2016), which was treated as a roll-out period. The broken line represents the commencement of CPOE implementation, with diamonds on the x-axis indicating the month of implementation for each individual ward; points are stacked where implementation occurred at multiple wards in the same month. In Figure B, points represent the total numbers of completed blood samples, centred at the date of CPOE implementation for each ward. Only those N=27 wards with at least 18 months of follow-up post-CPOE were included in the analysis. Trend lines are from a segmented regression model. CPOE – Computerised Provider Order Entry.

### Laboratory turnaround time (LTAT) analysis

Segmented regression analysis of LTAT only included the N=27 wards with at least 18 months of follow-up after the implementation of CPOE. For these wards, the average LTAT was 209.2 minutes (95% CI: 205.0 – 213.3) 24 months prior to CPOE implementation, and did not change significantly over the period prior to CPOE being implemented (p=0.623, **Table 1**, **Figure 3**). However, a step-change reduction in LTAT was observed immediately after the CPOE implementation, with a reduction in the median of 31.7 minutes (95% CI: 25.5 – 37.9, p<0.001). The median LTAT then remained stable over the 18 months post-CPOE, with no significant change in the gradient observed (p=0.085).

**Figure 3.**
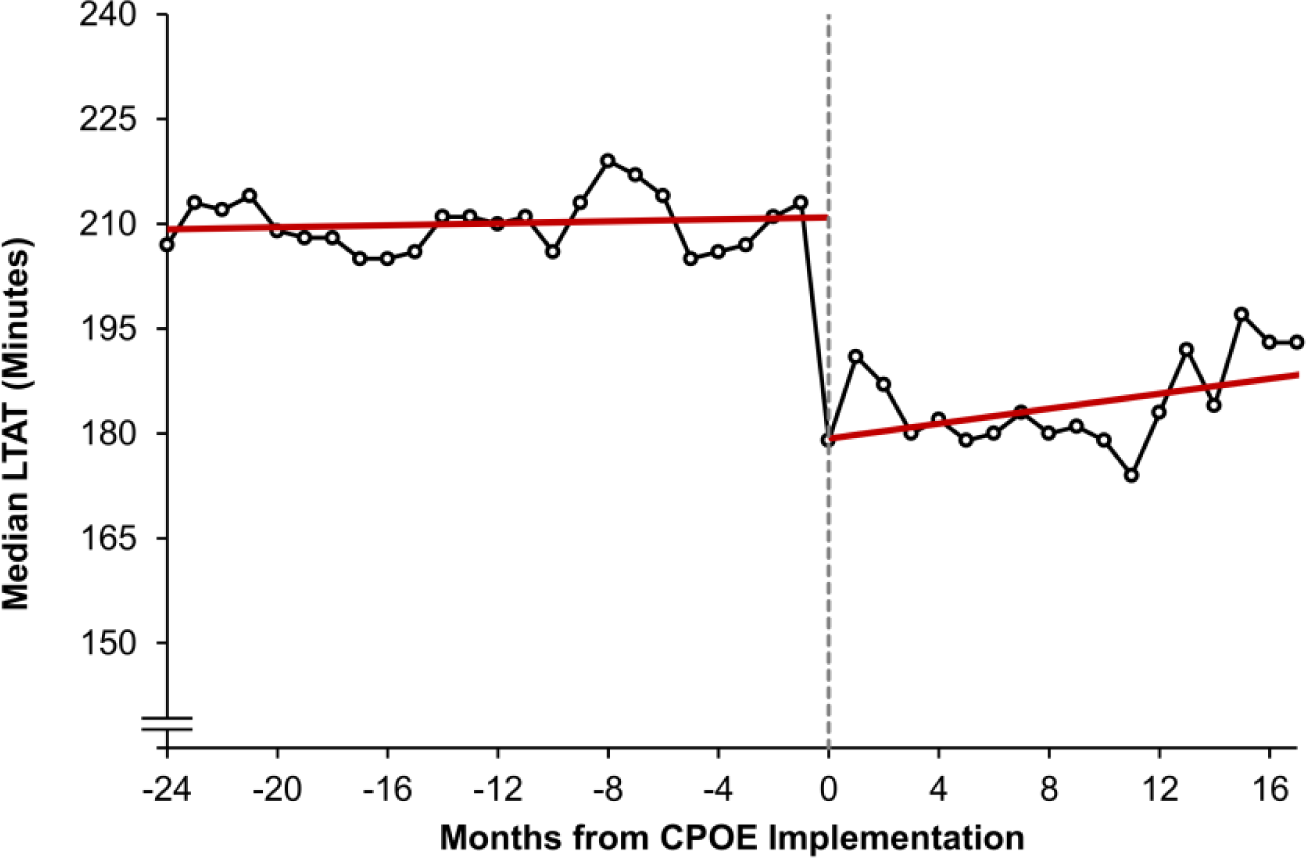
Segmented regression model of the median LTAT for inpatient blood samples. **Legend.** Points represent the median LTAT within each calendar month, with Month 0 (and the broken line) designating the month in which CPOE was implemented. Only those N=27 wards with at least 18 months of post-CPOE follow-up were included, to maintain a consistent cohort. Trend lines are from a segmented regression model, as described in Table 1. CPOE – Computerised Provider Order Entry, LTAT – Laboratory Turnaround Time.

**Table 1.** Segmented regression model of the median LTAT for inpatient blood samples Legend. : Results are from a segmented linear regression model of the monthly median LTAT over the 24 months pre- and 18 months post-CPOE implementation, as described in the Methods section. Only those N=27 wards with at least 18 months of post-CPOE follow-up were included, to maintain a consistent cohort. Coefficients are reported in minutes, and gradients are reported per calendar month. Bold p-values are significant at p<0.05. CPOE – Computerised Provider Order Entry, LTAT – Laboratory Turnaround Time.

|  | Coefficient<br>(Minutes; 95% CI) | p-Value |
| --- | --- | --- |
| Intercept | 209.2 (205.0, 213.3) | **<0.001** |
| Gradient Pre-CPOE (per Month) | 0.1 (-0.2, 0.4) | 0.623 |
| Step-Change Post-CPOE | -31.7 (-37.9, -25.5) | **<0.001** |
| Change in Gradient Post-CPOE (per Month) | 0.5 (-0.1, 1.0) | 0.085 |

To review whether changes in LTAT varied across clinical settings with the hospital, subgroup analyses were then performed within different groupings of wards (**Table 2** and **Figure 4**). Whilst there was variability in the impact of CPOE across these subgroups of wards, significant step-change improvements were still observed in the groups of medicine, surgery and non-critical care wards. CPOE was not found to be associated with a step-change in the median LTAT for the subgroup of critical care wards (p=0.185, **Figure 4a**). However, a significant reduction in the gradient (p<0.001) was instead seen, indicating that improvements in LTAT in the critical care wards were gradual, rather than occurring immediately post-CPOE implementation.

**Figure 4.**
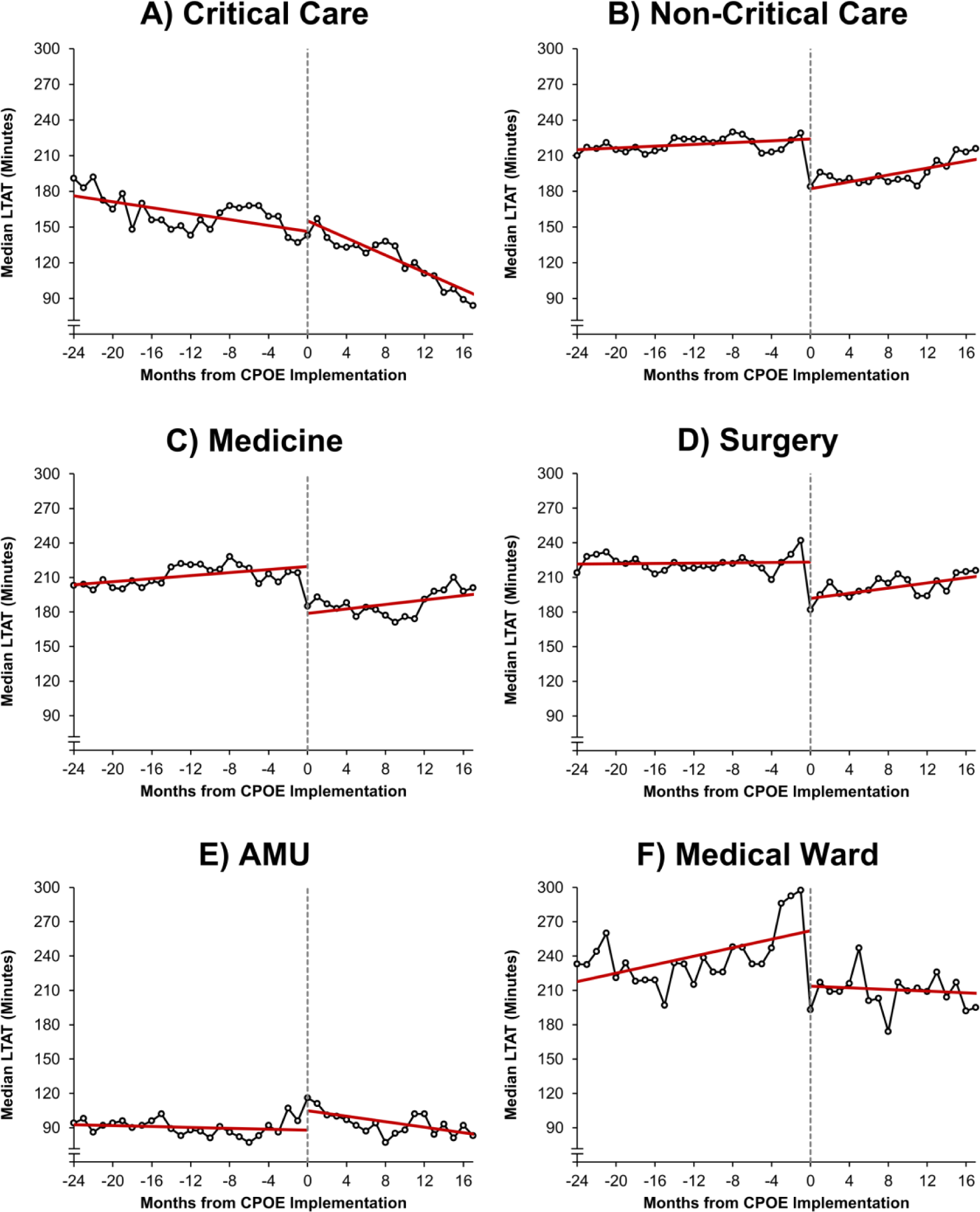
Segmented regression models of the median LTAT for inpatient blood samples by specialty/area Legend. Points represent the median LTAT within each calendar month, with Month 0 (and the broken line) designating the month in which CPOE was implemented. Figures A-D include only those wards with at least 18 months of follow-up post-CPOE implementation, to maintain a consistent cohort; Figures E-F represent data for individual wards. Trend lines are from a segmented regression model on the stated specialty/area, as described in Table 2. AMU - Acute Medical Unit, CPOE – Computerised Provider Order Entry, LTAT – Laboratory Turnaround Time.

**Table 2.** Segmented regression models of the median LTAT for inpatient blood samples by specialty/area Legend: Results are from segmented linear regression models of the monthly median LTAT times over the 24 months pre- and 18 months post-CPOE implementation, as described in the Methods section. Separate models were produced for each of the specialty groups/areas. Analyses of Group 1 and 2 include only those N=27 wards with at least 18 months of post-CPOE follow-up, to maintain a consistent cohort; analysis of Group 3 is based on individual wards. Coefficients are reported in minutes, and gradients are reported per calendar month. Bold p-values are significant at p<0.05. AMU – Acute Medical Unit, CPOE – Computerised Provider Order Entry, LTAT – Laboratory Turnaround Time.

|  | Coefficient<br>(Minutes; 95% CI) | p-Value | Coefficient<br>(Minutes; 95% CI) | p-Value |
| --- | --- | --- | --- | --- |
|  | **Group 1** |  |  |  |
|  | *Critical Care* |  | *Non-Critical Care* |  |
| Intercept | 177.5 (168.6, 186.4) | **<0.001** | 214.6 (209.3, 219.9) | **<0.001** |
| Gradient Pre-CPOE | -1.2 (-1.9, -0.6) | **<0.001** | 0.4 (0.0, 0.7) | **0.047** |
| Step-Change Post-CPOE | 8.9 (-4.4, 22.3) | 0.185 | -41.8 (-49.8, -33.9) | **<0.001** |
| Change in Gradient Post-CPOE | -2.4 (-3.5, -1.2) | **<0.001** | 1.1 (0.4, 1.8) | **0.003** |
|  | **Group 2** |  |  |  |
|  | *Medicine* |  | *Surgery* |  |
| Intercept | 203.0 (195.9, 210.2) | **<0.001** | 221.4 (215.2, 227.7) | **<0.001** |
| Gradient Pre-CPOE | 0.7 (0.2, 1.2) | **0.011** | 0.1 (-0.4, 0.5) | 0.744 |
| Step-Change Post-CPOE | -40.7 (-51.4, -30.0) | **<0.001** | -31.4 (-40.8, -22.0) | **<0.001** |
| Change in Gradient Post-CPOE | 0.3 (-0.6, 1.2) | 0.494 | 1.0 (0.2, 1.9) | **0.012** |
|  | **Group 3** |  |  |  |
|  | *AMU* |  | *Medical Ward* |  |
| Intercept | 92.8 (86.3, 99.4) | **<0.001** | 215.8 (199.6, 232.0) | **<0.001** |
| Gradient Pre-CPOE | -0.2 (-0.7, 0.3) | 0.381 | 1.9 (0.7, 3.0) | **0.002** |
| Step-Change Post-CPOE | 17.0 (7.2, 26.8) | **0.001** | -48.5 (-72.7, -24.2) | **<0.001** |
| Change in Gradient Post-CPOE | -1.0 (-1.8, -0.2) | **0.020** | -2.2 (-4.3, -0.1) | **0.038** |

Analysis of selected individual wards identified a different effect of CPOE implementation in one acute clinical setting, namely the Acute Medical Unit (an assessment unit for acutely unwell adults). Pre-CPOE, the LTAT at this ward was considerably shorter than the remainder of wards, with a median of 92.8 minutes (95% CI: 86.3 – 99.4), compared to 209.2 minutes (95% CI: 205.0 – 213.0) for the cohort as a whole. The implementation of CPOE within AMU was associated with a step-change increase in the median LTAT of 17.0 minutes (95% CI: 7.2 – 26.8, p=0.001, **Figure 4e**). However, this was followed by a reduction in LTAT (p=0.020), such that the median LTAT at 18 months post-CPOE implementation was lower than it had been immediately prior to CPOE implementation (77.1 vs. 87.9 minutes).

### Sub-section sensitivity analysis – Laboratory turnaround time (Lab-LTAT)

A sub-section analysis was completed, which only assessed the turnaround time within the laboratory, namely the Lab-LTAT. The median Lab-LTAT was found to show a greater improvement than the overall LTAT after the implementation of CPOE, with a step-change reduction of 52.5 minutes (95% CI: 47.9 – 57.0, p<0.001, **Table 3** and Figure 5). Supplementary Lab-LTAT analyses were conducted for the same clinical subgroups that were explored in **Table 2** (see: **Supplementary Table 1** and **Supplementary** Figure 2). These analyses consistently found reductions in Lab-LTAT compared to the analysis of the overall LTAT apart from the AMU, which the median Lab-LTAT remaining consistent over the whole study period for this ward (p=0.285).

**Figure 5.**
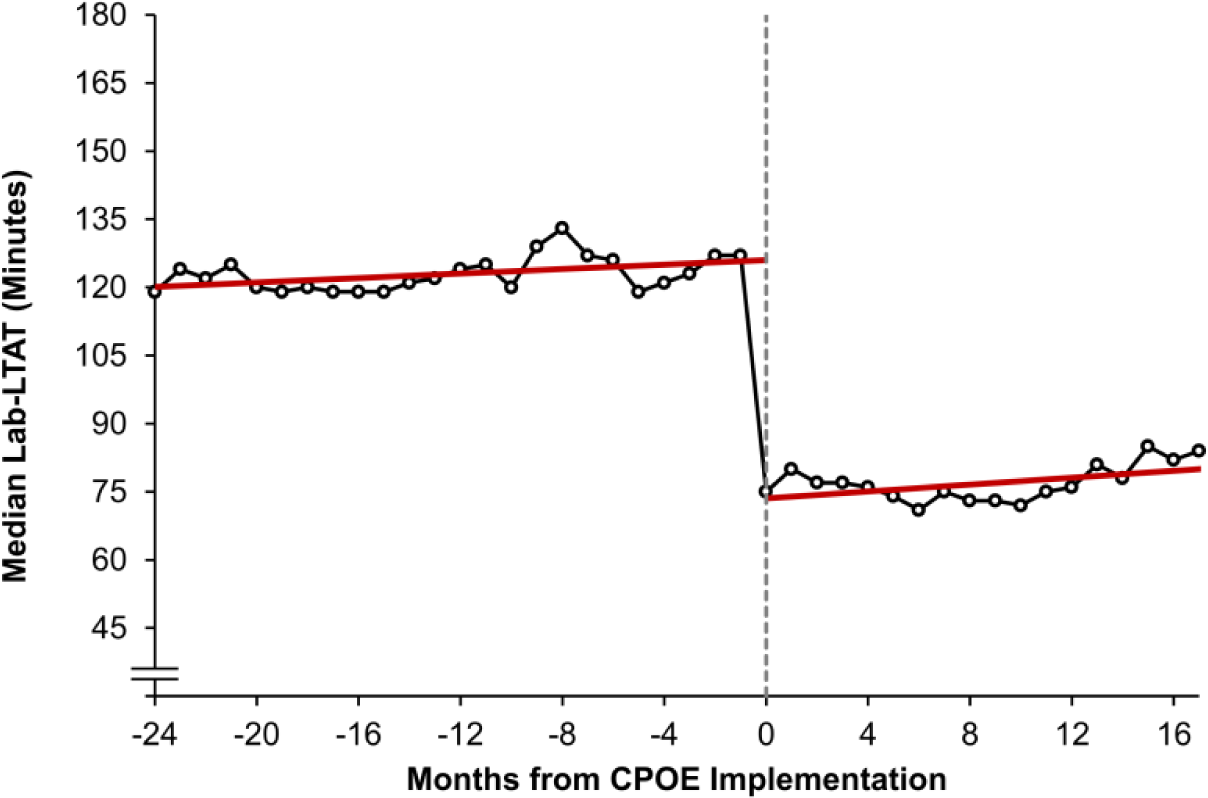
Segmented regression model of the median turnaround times for inpatient blood samples solely within the laboratory (Lab-LTAT). Legend. Points represent the median Lab-LTAT within each calendar month, with Month 0 (and the broken line) designating the month in which CPOE was implemented. Only those N=27 wards with at least 18 months of follow-up post-CPOE implementation were included, to maintain a consistent cohort. Trend lines are from a segmented regression model, as described in Table 3. CPOE – Computerised Provider Order Entry, LTAT – Laboratory Turnaround Time.

**Table 3.**
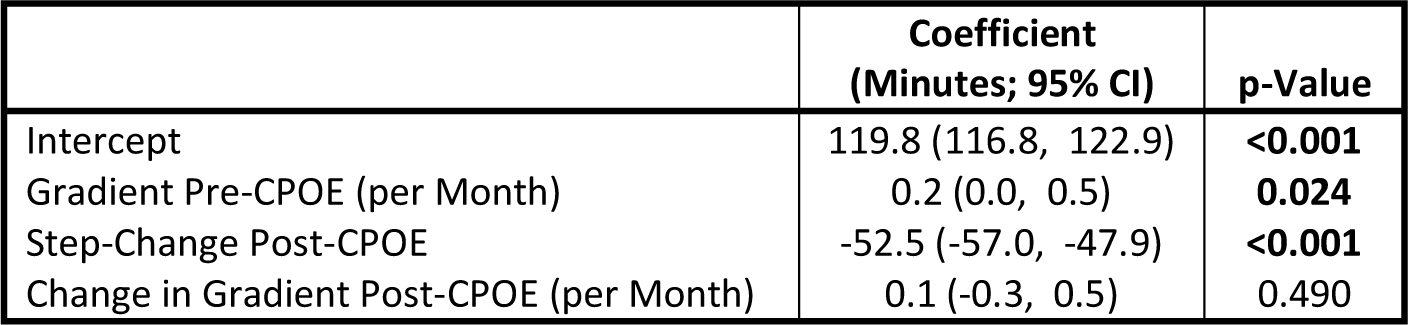
Segmented regression model of the median turnaround times for inpatient blood samples solely within the laboratory (Lab-LTAT). Legend. Results are from a segmented linear regression model of the monthly median Lab-LTAT over the 24 months pre- and 18 months post-CPOE, as described in the Methods section. Only those N=27 wards with at least 18 months of follow-up post-CPOE implementation were included, to maintain a consistent cohort. Coefficients are reported in minutes, and gradients are reported per calendar month. Bold p-values are significant at p<0.05. CPOE – Computerised Provider Order Entry, LTAT – Laboratory Turnaround Time.

|  | Coefficient<br>(Minutes; 95% CI) | p-Value |
| --- | --- | --- |
| Intercept | 119.8 (116.8, 122.9) | **<0.001** |
| Gradient Pre-CPOE (per Month) | 0.2 (0.0, 0.5) | **0.024** |
| Step-Change Post-CPOE | -52.5 (-57.0, -47.9) | **<0.001** |
| Change in Gradient Post-CPOE (per Month) | 0.1 (-0.3, 0.5) | 0.490 |

## DISCUSSION

To our knowledge, this is the first study documenting the change in LTAT after the implementation of CPOE for laboratory tests across all clinical areas within a whole system hospital setting. CPOE implementation was associated with an overall step-change reduction in the median LTAT of 31.7 minutes, which was sustained over the 18 months post implementation. Although this varied by broad clinical specialty, most clinical areas either demonstrated a step-change reduction in median LTAT or a change in the gradient of LTAT post-CPOE favouring faster reporting of results.

Potential reasons for the reduction in LTAT include the operational efficiency gains from replacing paper-based orders with computerised ordering systems. Barcodes interface with the CPOE system and eliminate the time previously taken to manually load and enter the sample data prior to commencing the analysis. The clinical impact of a reduction in LTAT has not been explored here; however, potential impacts may include faster clinical decision making where dependant on a laboratory result.

Subgroup analyses by specialty and ward generally found similar step-change improvements in LTAT after implementation of CPOE. However, no such improvement was observed for the Acute and Critical Care wards, with AMU demonstrating an increase in LTAT immediately after CPOE was introduced. New CPOE processes in AMU may have disrupted sample collection. This is in keeping with in other studies of CPOE in acute care settings which have reported increases in timed processes in Emergency Departments (ED) following the implementation of CPOE,^(14)^ with authors suggesting that the complexity of the acute environment makes them vulnerable to disruption following the introduction of any new service.^(17)^ However, in the current study there was a progressive improvement in LTAT after CPOE implementation in both AMU, and Critical Care. Consequently, the median LTAT at 18 months after CPOE implementation was lower than it had in the prior period suggesting benefit in the long-term.

As no other major changes to the conveyance, processing, reporting or quality assurance processes for blood samples occurred during the study period, the reduction in LTAT is most likely to reflect the impact of CPOE. The Lab-LTAT analysis indicates this element of the request cycle saw the greatest benefit of CPOE implementation. There is relatively little research into the impact of CPOE on hospital pathology services; however, this study suggests improvements in efficiencies.

An additional finding of this study was a significant increase in the number of samples processed following the implementation of CPOE to a ward, equivalent to an additional 12,541 samples per month across the hospital, representing a 31% increase. That the LTAT did not lengthen, and rather was shortened significantly, despite this substantial increase in sample processing suggests the benefits of CPOE are resilient to service change.

The reasons for, and implications of the significant increase in the number of samples following the implementation of CPOE were not explored here. A report reviewing the Diagnostic Services for NHS England^(18)^ and a Primary Care study^(19)^ over a comparable period described an increase in clinical test ordering, so this increase has been noted elsewhere in healthcare settings. It is unclear if the increase was causally associated with CPOE implementation. A potential reason for this may be the CPOE system making it easier for samples to be ordered in panels as well as individually.

A strength of this study was the large sample size and duration of the study, with the analysis of LTAT including data from almost two million samples collected over a six-year period. This gave sufficient statistical power to detect small changes in LTAT, and permitted subgroup analyses, both by specialty and for individual wards. However, the results must be interpreted in light of the limitations of the study. Whilst QEHB is a large hospital treating a diverse case mix of patients across a wide range of specialties, the results only represent the experience of a single centre. As such, the findings may not necessarily be generalisable to other centres, particularly those where processes and procedures differ considerably from those at QEHB. The study was also purely quantitative meaning that, whilst trends in LTAT could be assessed, no qualitative evidence was collected to examine the reasons for the observed changes in LTAT across the clinical settings.

## CONCLUSION

This study demonstrates that the implementation of CPOE within an acute hospital improves blood result reporting through reduced LTATs which were sustained over time despite a large increase in test ordering. LTAT in acute care settings were initially adversely affected, but did improve subsequently, suggesting that system changes in these complex settings needs careful consideration during deployment. Understanding the likely reduction in LTAT also provides information to support an economic evaluation of the implementation of such a system into a new setting.

## Data Availability

To facilitate knowledge in this area, the anonymised participant data and a data dictionary defining each field will be available to others through application to PIONEER via the corresponding author.

## Acknowledgements

This work was supported by PIONEER, the Health Data Research Hub in acute care and the HDR-UK Better Care programme. This work uses data provided by patients and collected by the NHS as part of their care and support. We would like to acknowledge the contribution of all staff, key workers, patients and the community who have supported our hospitals and the wider NHS.

The research was delivered as part of the NIHR Midlands Safety Research Collaboration, NIHR Midlands Applied Research Collaborative and NIHR Birmingham Biomedical Research Centre.

## Conflicts of Interest

X. Zou, F. Evison, J. Hodson, J. Atia, C. Webster, M. Garrick, J. Coleman, T Pankhurst report no conflicts of interest. S Gallier reports funding support from HDRUK, MRC and NIHR. S Ball reports funding support from HDRUK. K. Nirantharakumar reports funding support from HDRUK and NIHR. E Sapey reports funding support from HDRUK, MRC, Wellcome Trust, NIHR, Alpha 1 Foundation, EPSRC and British Lung Foundation.

## Author contribution

S Gallier, X Zou, J. Hodson, F Evison, J Atia, J Coleman, T Pankhurst, E Sapey designed the study, collated data, performed some analysis, wrote the manuscript. X Zou, J Hodson and F Evison curated data and supported statistical analysis. S Gallier performed statistical analysis and wrote the first draft of manuscript. M Garrick supported data collection. All authors amended the manuscript and approved the final version.

## Notes

### Funding Statement

This study did not receive any funding.

### Author Declarations

The study was supported by PIONEER, a Health Data Research Hub in Acute Care. Ethical approvals for the study were provided by the East Midlands Derby REC (reference: 20/EM/0158).

