## Supplementary Figure 1 for "The Impact of Electronic Order Communications on Laboratory Turnaround Times in Acute Hospital Care"

### SUPPLEMENTARY FIGURES

**Supplementary Figure 1 – Modified Consort flow diagram explaining how the eligible samples for analyses were selected.**

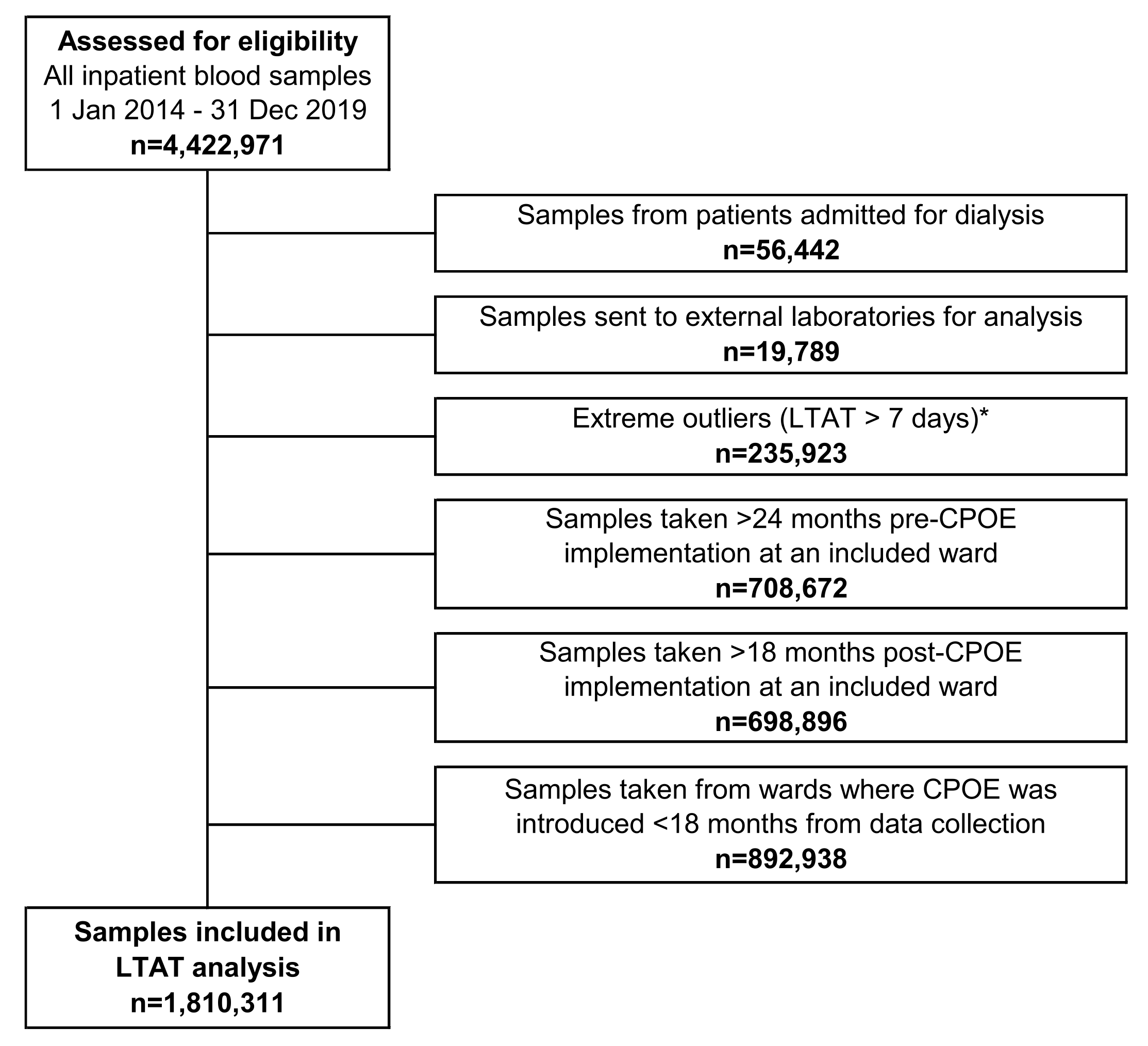

**Legend.** **Samples were defined as extreme outliers if the results were not returned within 7 days (10,080 minutes); the majority of such cases represented either instances where tests were ordered but no sample was taken, or highly specialised tests that required considerable time to be analysed, both of which were outside the scope of this study. CPOE – Computerised Provider Order Entry, LTAT – Laboratory Turnaround Time.*
