## Supplementary Figure 2 for "The Impact of Electronic Order Communications on Laboratory Turnaround Times in Acute Hospital Care"

### SUPPLEMENTARY FIGURES

**Supplementary Figure 2 – Segmented regression models of the median Lab-LTAT for inpatient blood samples by specialty/area**

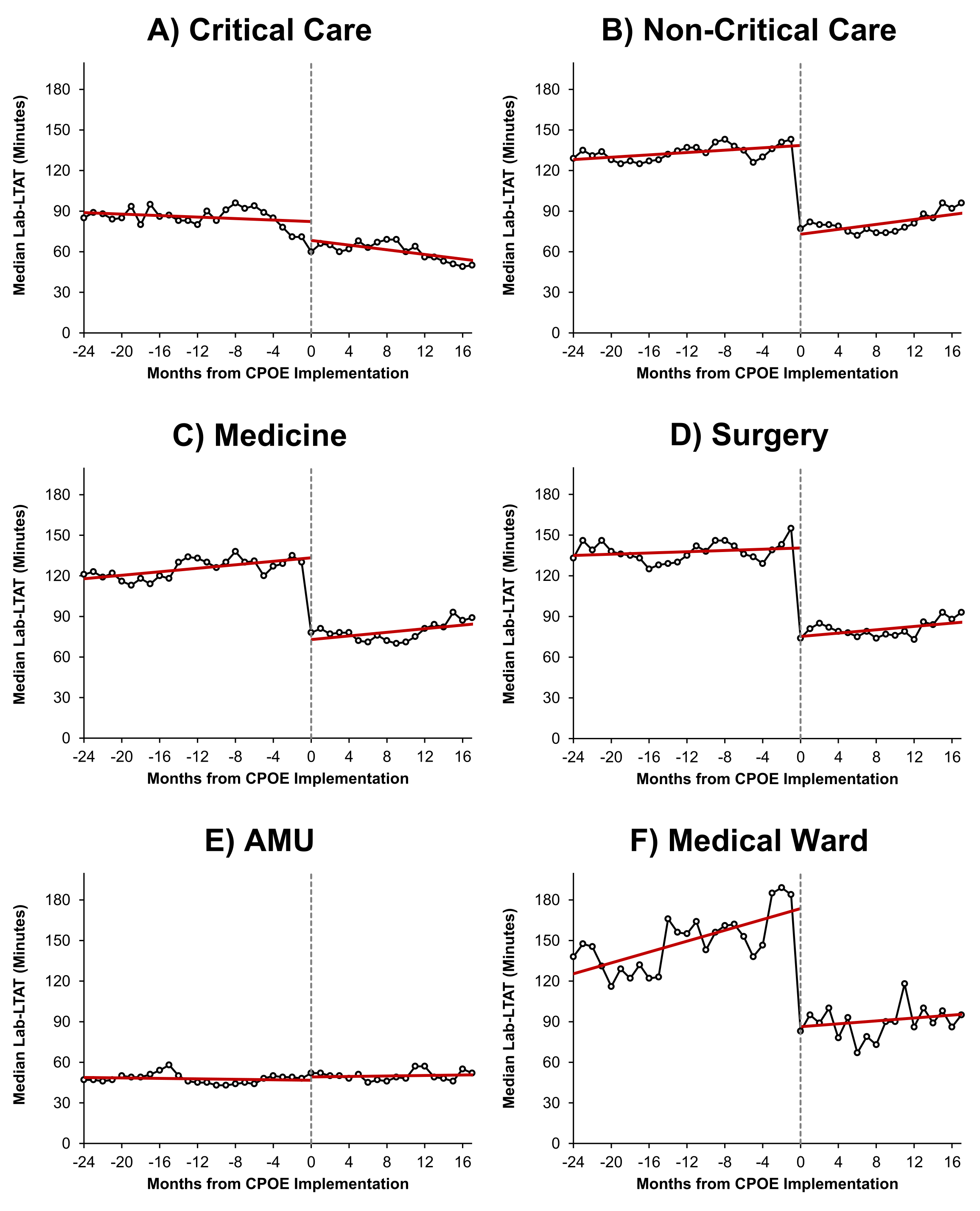

**Legend***. Points represent the median Lab-LTAT within each calendar month, with Month 0 (and the broken line) designating the month in which CPOE was implemented. Figure A-D include only those wards with at least 18 months of post-CPOE follow-up, in order to maintain a consistent cohort; Figure E-F represent data for individual words. Trend lines are from a segmented regression model on the stated specialty/area, as described in* ***Supplementary Table 1****. CPOE – Computerised Provider Order Entry, LTAT – Laboratory Turnaround Time.*
